# Glucose response to low-dose glucagon for insulin-induced mild hypoglycaemia after 12 weeks of high versus low carbohydrate diet. A pilot study

**DOI:** 10.1101/2020.10.30.20222794

**Authors:** Ajenthen G. Ranjan, Signe Schmidt, Jens J. Holst, Merete B. Christensen, Kirsten Nørgaard

## Abstract

**Objective:** To compare the glucose response to low-dose glucagon after high carbohydrate diet (HCD) versus low carbohydrate diet (LCD).

**Methods:** Individuals with insulin pump-treated type 1 diabetes went through 12 weeks of HCD (>250 g/day) and 12 weeks of LCD (<100 g/day) in random order and separated by 12 weeks. At end of each diet, mild hypoglycaemia was induced in the fasting state by a subcutaneous insulin bolus. When plasma glucose (PG) reached 3.9 mmol/L, 100 µg glucagon was given subcutaneously.

**Results:** Four of six participants completed both study visits while the remaining two only completed the study visit following LCD. They were 37 (28-52) years old (median (IQR)), had BMI 25.0 (24.5-25.2) kg/m^2^, and HbA1c 57 (55-59) mmol/mol or 7.4 (7.2-7.5) %. Daily carbohydrate intake was 95 (86-97) g during LCD and 254 (184-259) g during HCD. Compared with HCD, LCD had a significantly lower area under the PG curve from 0-120 min (521 (394-617) vs 663 (546-746) mmol/l x min, p=0.045) and insignificant lower incremental PG peak after the glucagon bolus (1.5 (0.6-3.2) vs 3.0 (2.2-4.2) mmol/L, p=0.317).

**Conclusion:** In conclusion, the glucose response to low-dose glucagon was reduced after 12 weeks of LCD compared with HCD.

## Introduction

Managing type 1 diabetes is challenging (1). Although patients strive for optimal glycaemic control, they are limited by the risk of mild and severe hypoglycaemia associated with insulin therapy (2). Low-doses of glucagon have shown effective in treating mild hypoglycaemia and may have the potential to serve as an alternative to oral glucose in these situations if patients are trying to avoid extra calorie intake and weight gain (3,4). We have previously shown that one week of restricted carbohydrate intake impaired the ability of glucagon to treat mild hypoglycaemia (5). In this study, we tested whether the reduction in the glucose response to glucagon persisted after 12 weeks of low carbohydrate diet (LCD). This pilot study is an extension of our previously published study that examined the glycaemic effects and risk markers of cardiovascular disease of the LCD and HCD (6).

## Materials and methods

### Design

This was a randomized open-labelled 2-arm crossover study. The interventions period had a duration of 12 weeks and were separated by a 12-week interval. We recruited adults (>18 years) with type 1 diabetes duration > 3 years, insulin pump treatment > 1 year, glycated haemoglobin (HbA_1c_) > 53 mmol/mol (>7.0%), BMI 20–27 kg/m^2^, who were practicing carbohydrate counting.

Key exclusion criteria were celiac disease, inflammatory bowel disease, and use of drugs other than insulin affecting glucose metabolism. Patients completed a screening visit, an insulin pump optimisation period and two randomly ordered dietary periods ending with a study visit. This pilot study focuses on these study visits and the results have not been reported before. The study procedures preceding these study visits and the outcomes have been described in detail elsewhere (6).

In brief, participants went through a screening and two 12-week diet periods, each ending with a study visit. After the screening a dietitian estimated each participant’s daily calorie intake based on their daily activity level, gender, age, height and weight as well as a 5-day pre-randomisation diet recording. Subsequently, isocaloric diets with high carbohydrate (>250 g/day) or low carbohydrate (<100 g/day) content were designed for each participant. The carbohydrate limits were decided based on experience from our outpatient clinic. Participants’ insulin pump settings were optimized both 2-3 weeks before and 2-3 weeks after each diet period start. The actual carbohydrate consumption during the intervention periods was assessed by the amount of carbohydrate that was prospectively registered by the participants in the insulin pumps.

Each 12-week diet period ended with an in-clinic study visit (7). During the 24 h before each study visit, participants avoided hypoglycaemia (continuous glucose monitor or capillary meter glucose ≤ 3.9 mmol/L), exercise, and alcohol consumption. They arrived fasting in the morning aiming for PG level of 7.0 mmol/L. Based on their arrival PG level and insulin-correction factor, an insulin bolus (NovoRapid, Novo Nordisk, Bagsværd, Denmark) was administered through their insulin pumps aiming to lower plasma glucose to 3.0 mmol/L. Once plasma glucose reached 3.9 mmol/L, a subcutaneous bolus of 100 µg glucagon (GlucaGen, Novo Nordisk, Bagsværd, Denmark) was administered (figure 1). Plasma glucose, plasma glucagon, serum insulin aspart, hypoglycaemic symptoms (Edinburgh Hypoglycaemia Scale), and visual analogue scales for side effects to glucagon were measured regularly during the visit.

**Figure 1:**
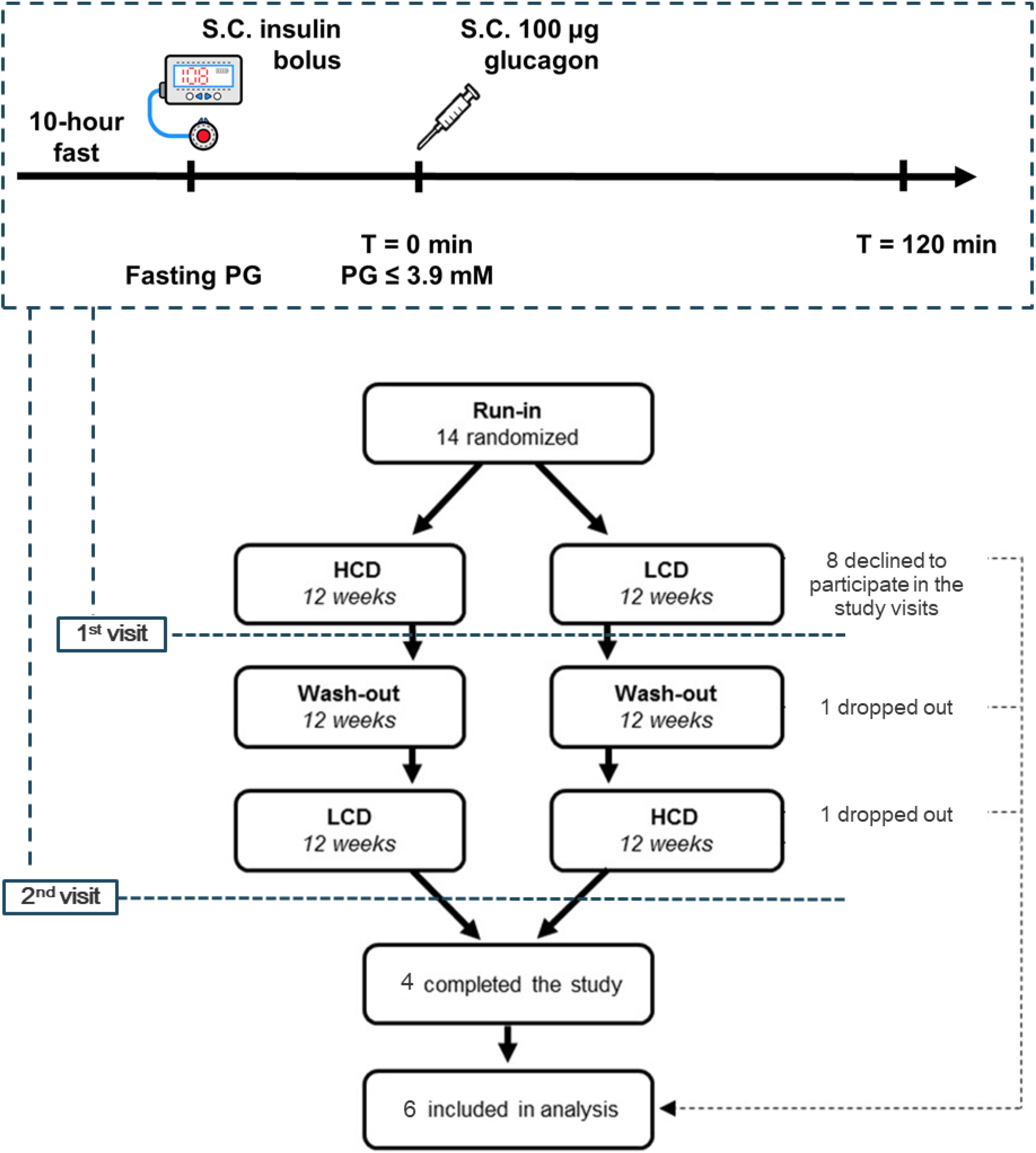
Overview of the study and the participant flowchart. Study participants are a subpopulation from another study (6). The study visits were performed after 12 weeks of high carbohydrate diet (HCD > 250 g) and after 12 weeks of low carbohydrate diet (LCD <100 g). Participants received a subcutaneous (S.C.) insulin bolus after 10 hours fasting. Once plasma glucose (PG) was below 3.9 mmol/l, a S.C. 100 µg glucagon bolus was given. Participants were observed for another two hours after the glucagon bolus.

In the main study, ten participants completed the LCD and HCD periods. In the present study, only a subpopulation of participants who consented (written and orally) to perform the subsequent study visits were included (figure 1).

The outcomes of interest were the positive incremental area under the curve from 0 to 120 min (AUC_0–120_), incremental PG peak from 0 to 120 min, and time-to-peak after the glucagon injection. An intention-to-treat analysis was performed. Friedman’s nonparametric analysis of variance for repeated measurements was used to compare the data after HCD and LCD. If data, however, were normally distributed, a linear mixed effect model was used with covariates including diet intervention (HCD and LCD), time, and intervention by time interaction as fixed-effects and subject as random effect. SAS 9.4 (SAS Institute, Inc., Cary, NC) was used. Based on sample size of 4 participants, the mean and standard deviation of difference in plasma glucose peak, the study has a power of 65%. We considered P<0.05 as statistically significant. Data are presented as median (interquartile range) in the text and as mean±sem in figures unless otherwise stated.

The Regional Committee on Health Research Ethics (H-1509662) and the Danish Data Protection Agency approved the study, which was conducted in accordance with the Helsinki Declaration. Clinicaltrial.gov: NCT02888691.

## Results

### Baseline

Four of six participants completed both study visits while the remaining two only completed the study visit after LCD (Figure 1). They were 37 (28-52) years old (median (IQR)) with BMI 25.0 (24.5-25.2) kg/m^2^ and HbA1c 57 (55-59) mmol/mol or 7.4 (7.2-7.5) %. The daily carbohydrate intake according to insulin pump recordings during each period was 95 (86-97) g and 254 (184-259) g, respectively.

### Study visits

The insulin dose to induce mild hypoglycaemia did not differ between the study visit after HCD and LCD (3.7 (2.05-6.10) and 4.5 (2.9-5.2) IU, p= 0.89). Plasma glucose levels were also similar between the two study visits at time of insulin administration (8.2 (7.2-14.9) mmol/l and 8.4 (7.6-9.8) mmol/l, p=0.32) and glucagon administration (3.62 (3.48-3.72) and 3.69 (3.60-3.75) mmol/l, p = 0.32). Plasma glucose increased to an incremental peak of 1.5 (0.6-3.2) mmol/l after LCD compared with 3.0 (2.2-4.2) mmol/L after HCD, p =0.31. The time to reach glucose peak after glucagon administration was the same in the two study visits (32.5 (27.5-40.0) and 35.0 (35.0-35.0) min, p=0.56). Glucagon restored euglycaemia (>3.9 mmol/l) within 10 (10-10) min after HCD and within 12 (10-15) min after LCD (p=0.16). The area under the glucose curve from 0-120 min (AUC_0-120_) after glucagon administration was, however, lower after LCD compared with HCD (521 (394-617) vs 663 (546-746) mmol/l x min, p=0.045) (figure 2).

**Figure 2:**
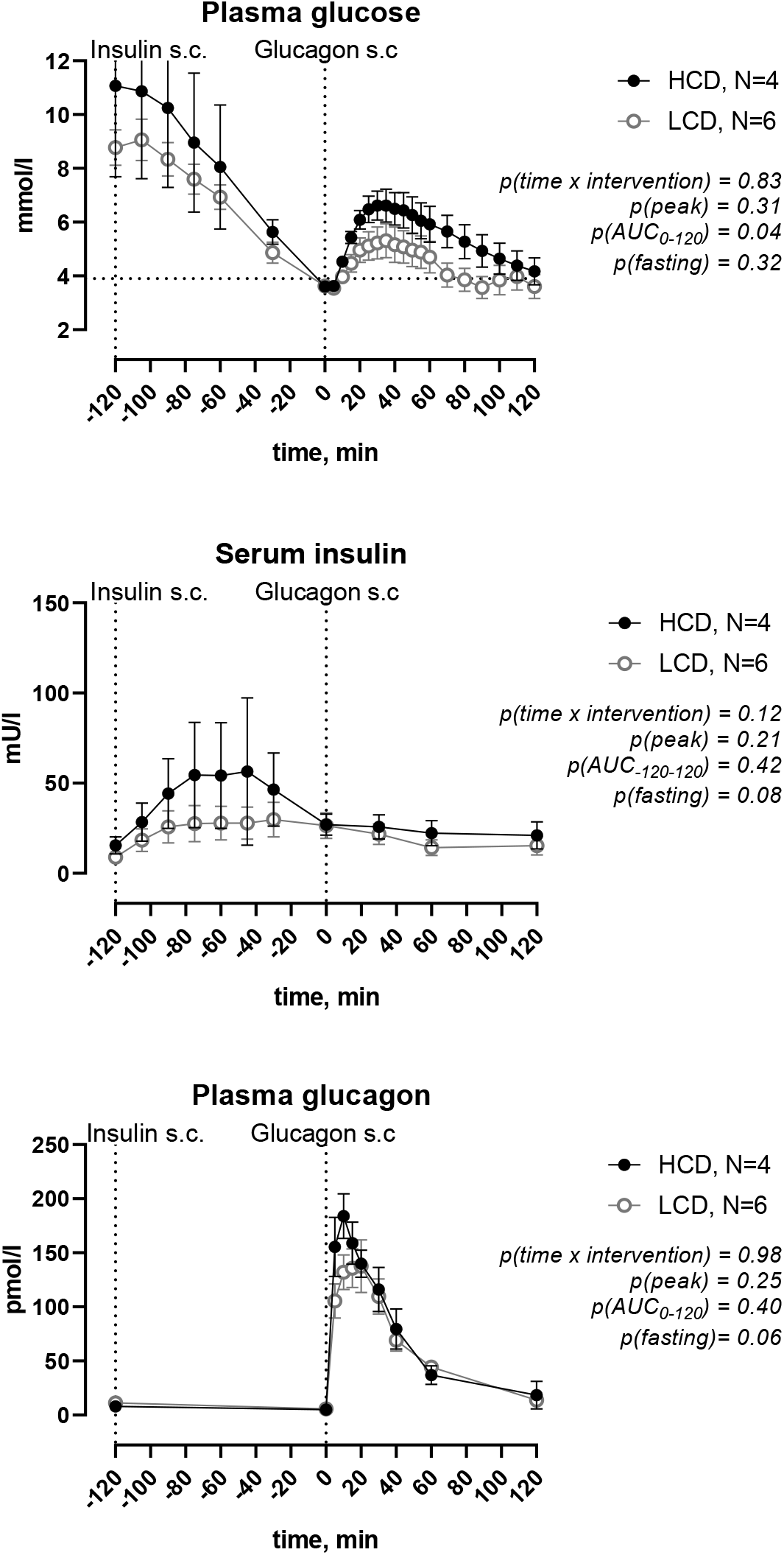
Mean±SEM concentrations of plasma glucose, serum insulin and plasma glucagon during the study visit after 12 weeks of high carbohydrate diet (HCD >250 g) and after 12 weeks of low carbohydrate diet (LCD < 100 g). Participants received a subcutaneous (s.c.) insulin bolus in the fasting state. Once plasma glucose was below 3.9 mmol/l, a s.c. 100 µg glucagon bolus was given. Participants were observed for two hours after the glucagon bolus. A repeated measurement ANOVA was used to compare the glucose, insulin and glucagon profiles between the study visits. P-value < 0.05 was considered significant.

### Insulin

Fasting serum insulin tended to be lower on LCD compared with HCD (8.3 (4.5-12.9) and 15.8 (7.2-23.9) mU/l, p=0.08). Peak serum insulin after insulin administration was similar between study days (LCD 20.1 (16-75.6) and HCD 45.3 (19.2-139.9) mU/l, p=0.21). Area under the curve for serum insulin from glucagon administration to end of study was similar between the study days (LCD 3110 (2774-7306) and HCD 5333 (2794-11224) mU/l x min; p=0.42) (figure 2).

### Glucagon

Fasting plasma glucagon tended to be higher on LCD compared with HCD (11.0 (8.0- 12.0) and 8.0 (4.0-12.0) pmol/l, p=0.055). Peak plasma glucagon after glucagon administration was similar between study days (LCD 154 (112-228) and HCD 198 (172-238) pmol/l, p=0.25). Area under the curve for plasma glucagon from 0-120 min after glucagon administration was similar between the study days (LCD 6960 (5355-8840) and HCD 7820 (6413-9473) pmol/l x min; p=0.40 (figure 2).

### Side effects

No differences in side effects, change in blood pressure or pulse as well as cognitive function during hypoglycaemia were seen between study visits.

## Conclusion

In persons with type 1 diabetes, the overall glucose response to low-dose glucagon, shown as AUC_0-120_, was significantly lower after 12 weeks of LCD compared with after 12 weeks of HCD. However, the incremental glucose peak after glucagon injection did not differ significantly after LCD compared with HCD. The insulin dose to induce mild hypoglycaemia and the plasma glucose at time of glucagon injection as well as the insulin and glucagon levels were similar during both study visits.

Overall, these findings are consistent with our previous study that investigated the glucose response to low-dose glucagon after one week of LCD and HCD (5). In the previous one-week and the present twelve-week study, 100 µg glucagon resulted in a 50 % reduced incremental plasma glucose peak during LCD compared with HCD. Yet, in this study the difference did not reach statistical significance which may have several explanations. First, the number of participants is much lower, underpowering the present study. Second, LCD in the one-week study consisted of 50 g carbohydrates per day compared with the present study where LCD was 100 g carbohydrates per day. The two-fold difference in daily carbohydrate intake between LCDs in the two studies may have affected the glucose response to glucagon. We chose 100 g carbohydrate as the lower limit due to the significant weight loss, discomfort and difficulties to adhere to the carbohydrate restriction in the one-week study. Furthermore, many of our patients in the outpatient clinic consume 100 g carbohydrate daily. Yet, peak incremental plasma glucose and AUC_0-120_ were reasonably comparable between the LCD period after one week and after 12 weeks. Third, we performed this study because we speculated that the hepatic glucose response to glucagon may have been restored due to adaption to LCD after a longer period. The fasting glucagon levels after LCD were significantly higher than after HCD and these high glucagon levels could potentially downregulate the glucagon receptors and reduce the glucagon sensitivity. Despite, the non-significant difference in incremental plasma glucose peak, the overall glucose response determined as AUC_0-120_ was clearly diminished after LCD compared with HCD in the present study and this despite a tendency toward elevated insulin levels on HCD prior to the glucagon administration.

In summary, this study shows that the glucose response to low-dose glucagon is diminished after 12 weeks of LCD. Therefore, acute and prolonged restriction of carbohydrate intake should be accounted for, when individuals with type 1 diabetes choose to treat mild hypoglycaemia by injecting low-dose glucagon.

## Data Availability

Datasets and the full trial protocol can be requested from the corresponding author.

## Acknowledgement

We thank the study participants for their time and effort. This work was funded by a research grant from the Danish Diabetes Association and the Danish Diabetes Academy supported by the Novo Nordisk Foundation.

## Conflict of interest

A.R. and M.B.C have nothing to disclose. S.S. has served on advisory boards for Roche Diabetes Care and Medtronic. K.N. serves as adviser to Medtronic, Abbott, Sanofi and Novo Nordisk, owns shares in Novo Nordisk, has received research grants from Novo Nordisk, Roche Diabetes Care and Zealand Pharma, and has received fees for speaking from Medtronic, Roche Diabetes Care, Rubin Medical, Sanofi, Novo Nordisk A/S, Zealand Pharma and Bayer. J.J.H. has served on advisory boards for Novo Nordisk.

## Author contributions

A.R. contributed to the study design, data collection, statistical analyses and data interpretation, and wrote and edited the manuscript. M.B.C., S.S., J.J.H and K.N. contributed to the study design, data collection, statistical analyses and data interpretation, and reviewed and edited the manuscript. A.R. and K.N. is the guarantor of this work and, as such, had full access to all the data in the study and takes responsibility for the integrity of the data and the accuracy of the data analysis.

## Notes

### Clinical Trial

NCT02888691

### Author Declarations

The Regional Committee on Health Research Ethics (H-1509662) and the Danish Data Protection Agency approved the study, which was conducted in accordance with the Helsinki Declaration.

